# Clinicodemographic Prediction of Overall Survival in Patients with Head and Neck Merkel Cell Carcinoma: A Machine Learning Approach

**DOI:** 10.1101/2025.04.05.25325298

**Authors:** Jehad A. Yasin, Abd-alrahman Obeid, Ramez M. Odat, Fares A. Qtaishat, Mohammad-Amer A. Tamimi, Osama M. Younis, Hanan M. Qasem, Nicole Issi, Yousef A. Ateiwi, Sakher Alshwayyat

## Abstract

**Background:** Merkel cell carcinoma (MCC) is a rare cutaneous neuroendocrine malignancy with a higher case-fatality rate than melanoma. The prognosis of MCC is complex and depends on many factors. In this study, we aimed to develop predictive survival models using machine learning (ML) algorithms and statistical techniques for patients with head and neck MCC.

**Methodology:** Using a cohort of 1,372 patients diagnosed with MCC of the head and neck region in the United States between 2000 to 2019 sourced from the Surveillance, Epidemiology, and End Results (SEER) Program, we developed and evaluated a cox-proportional hazards (CPH) regression model, nine classification and regression ML models, and two ML-based survival models. Models were built with a total of 20 features, including demographic, cancer-related and treatment/surgery-related variables. Pre-processing, hyperparameter tuning, classification, regression, survival analyses, and model evaluations were performed using ‘scikit-learn’, ‘scikit-survival’, and ‘lifelines’ packages on Python 3.80.

**Results:** The mean age of patients was (66.0). Most cases were diagnosed in stage I (n=723, 52.7%). Multivariate CPH model yielded a Concordance-Index (C-Index) = 0.700 on the test set, outperforming both random forest survival (C-Index = 0.591) and survival tree (C-Index = 0.582) algorithms. Of the nine classification models, gradient boosting classifier predicted the most accurate 2-year (AUC = 0.75; accuracy= 0.71) and 5-year (AUC = 0.75; accuracy = 0.68) survival. Additionally, the ridge- and lasso-regularized linear models performed the most accurate regression (RMSE = 1182.84, R2 = 0.2259; RMSE = 1184.70, R2 = 0.2234, respectively), and the gradient boosting regressor had acceptable predictions (RMSE = 1189.64, R2 = 0.2170) on test sets. According to the Shapley Additive Explanations (SHAP) value analysis, the most critical feature of these regression models was age, followed by sex and AJCC stage.

**Conclusions:** This study found that machine learning and statistical models provide reliable survival predictions for head and neck Merkel cell carcinoma, with models like gradient boosting classifiers having acceptable outputs, especially for 2-year survival.

## 1.0 Introduction

Merkel cell carcinoma (MCC) is a rare and aggressive form of cutaneous malignancy that arises from Merkel cells, specialized neuroendocrine cells found in the epidermis (1, 2). MCC is predominantly affecting people older than 65 years (3). It is characterized by frequent regional and distant metastases, high recurrence rates, and lower overall survival compared to melanoma (4, 5). Previous studies estimate that the number of MCC cases will rise to 3,284 annually by 2025, with a significant increase expected among individuals aged ≥ 65 years (6). In the United States, the incidence has escalated from 0.15 cases per 100,000 in 1986 to 0.7 cases per 100,000 in 2016 (7). This surge is attributed to factors such as an aging population, increased UV exposure, and a growing number of immunosuppressed individuals (8). Early detection and prompt treatment are essential to mitigate the high morbidity and mortality associated with MCC (9).

Patients with MCC of head and neck region usually have poor prognosis with a five-year OS was 49.8% for stage I to 18.5% for stage IV (10). Around 50% of MCC cases occur in the head and neck region, where treatment is particularly challenging due to difficulties in achieving wide surgical margins and the complex lymphatic drainage in this area. Head and Neck Merkel Cell Carcinoma (HNMCC) presents distinct clinical challenges due to its high risk of local relapse, even in early stages, particularly in facial regions (5). Also, survival outcomes can vary significantly, depending on several key factors including tumor stage, presence of molecular aberrations present, feasibility of surgical resection, and the presence of underlying diseases (11, 12). Patients with HNMCC exhibit poorer survival compared to those with MCC in other regions, likely due to factors such as narrow surgical margins, unpredictable metastatic behavior, and tumor-specific characteristics; mortality is predominantly driven by distant metastases (13).

Helping to improve these processes, the field of machine learning (ML) has emerged as a promising tool in recent years, using routinely collected clinical and demographic data to predict various outcomes. The management of head and neck MCC requires early, accurate diagnosis and a multidisciplinary approach, incorporating surgery, radiotherapy, and, where indicated, chemotherapy or even immunotherapy (13, 14). Recent advancements in artificial intelligence (AI) are rapidly transforming the healthcare industry by increasing diagnostic accuracy, streamlining personalized plans, and improving patient outcomes across various medical fields by enhancing early disease detection (15). The integration of AI into diagnostic medicine has introduced new opportunities for precision in oncologic pathology, with applications showing a clear impact on improving survival outcomes. Machine learning model cohort study has also shown promise in identifying intermediate-risk head and neck squamous cell carcinomas patients who may benefit from specific therapies, such as chemoradiation treatments (16).

HNMCC presents distinct challenges due to its unique prognostic markers, emphasizing the importance of conducting focused future studies to accurately identify these markers and assess treatment outcomes (17).ML has shown promise in improving the prediction of prognosis, known to be particularly challenging in MCC of head and neck region (18, 19). The treatment for MCC varies depending on the stage of the disease. In the early stages, treatment typically involves resection and radiotherapy, whereas metastatic disease requires a combination of surgery, radiation therapy, and systemic therapy. Emerging data have shown improved outcomes with programmed cell death protein 1 axis agents (20). Systemic therapy, including immune checkpoint inhibitors, has become a mainstay for metastatic or locally advanced MCC, demonstrating improved overall survival, progression-free survival, and potential for durable responses in some patients (9, 21). Although surgical intervention remains crucial for localized tumors, the addition of adjuvant radiotherapy may only cure a fraction of patients (20).

ML models outperform traditional single-factor evaluations, helping identify patients for aggressive treatments while reducing unnecessary interventions, enabling better palliative care and symptom management (22, 23). Using ML and predictive modeling modalities, our study aimed to predict overall survival in MCC of head and neck region through using of a comprehensive set of demographics and clinicopathological variables derived from a large real-world data. Alongside highlighting several benefits of model integration in clinical decision-making, we also consider the challenges associated with their use.

## 2.0 Methods

### Data Collection and Processing

Data for this study was sourced from the The Surveillance, Epidemiology, and End Results (SEER) program (24). We accessed data consisting of demographic and clinicopathological profiles of patients with Head and Neck Merkel Cell Carcinoma. Processing of raw data initially involved cleaning by excluding empty/Not Applicable entries in addition to irrelevant variables. Data cleaning was followed by coding to express categorical data as binary vectors, with a label-encoding approach used for ordinal data (such as tumor stage) and one-hot-encoding for nominal inputs, converting them into several new columns. Python packages like “pandas” and “numpy” were essential for data manipulation, preprocessing, and performing complex mathematical operations on multi-dimensional arrays. A total of 1372 patients were included in the final analysis. Variables related to age group, sex, race, marital status, TNM and AJCC stages, surgery, primary tumor site, laterality, and type of therapy and surgery were included. Of the 32 extracted predictive features, some binary-hot encoded ones were excluded to avoid redundancy and collinearity in data, ending up with a total of 25 predictive features. Some variables, such as receiving chemotherapy or radiotherapy, included ambiguous responses labeled as "Yes" or "No/Unknown." For the purpose of analysis, we treated "No/Unknown" as "No." Afterwards, the processed dataset was randomly split into training and testing with 80:20 ratio.

### Statistical Data Analysis

Descriptive statistics were computed to summarize the key characteristics of patients. The multivariate cox proportional hazards regression (CPH) model was computed for potential predictors of survival. Hazard ratios (HRs) with 95% confidence intervals were also reported, and a forest plot was visualized using “matplotlib” along with “seaborn” on Python 3.80 (25, 26). Spearman’s correlations, Mann-Whitney U tests and Kaplan Meier analyses were done using IBM Statistical Package for the Social Sciences (SPSS) version 27 (27). For all statistical tests used, a p-value less than 0.05 was deemed statistically significant.

### Machine Learning (ML) Model Development and Testing

A wide array of computational tools were used for ML Models development using Python, with libraries such as “scikit-learn” for most machine learning models (28), “TensorFlow” for deep learning-based models (29).

For the prediction of 2-year and 5-year binary survival outcomes, we employed multiple ML classification models, including K-Nearest Neighbors (KNN), Multilayer Perceptron (MLP), Support Vector Machines (SVM), Logistic Regression (LR), Decision Trees (DT), Random Forest (RF), Gradient Boosting (GB), AdaBoost (ADA), and Gaussian Naive Bayes (GNB). The MLP models were implemented with two architectures: MLP-S, using scikit-learn, and MLP-TF, a 2-hidden layer (*64 and 32 nodes for first and second dense layers, respectively*) model using TensorFlow. Regression models for survival months were also constructed, such models included Unregularized and Lasso/Ridge-Regularized linear regression, MLP regressor, gradient boosting regressor, and random forest regressor. Specialized survival models like random forest survival (RFS) and survival decision trees.

Using the training set (80% of the entire dataset), the models were trained by iteratively adjusting model weights and parameters. For hyperparameter tuning, a Grid Search was conducted for each model using “GridSearchCV” library, which was done within a five-fold cross validation framework. This helps in identification of the best parameters for each model (for instance, number of neighbors in KNN, depth of the trees in Decision Trees and Random Forest and learning rates in boosting methods). MLP-S constituted two different models, MLPS2 and MLPS5, both using scikit-learn but emerged with different optimal hyperparameters. Best parameters for classification models built using scikit-learn are summarized in **Supplementary Table 1**.

Using the testing dataset (20% of the total dataset), the model performances were evaluated using metrics such as F1-Score, Accuracy, Precision, Recall, Area Under the Receiver Operating Characteristic (AUROC), and C-Index. ROC plots were generated for each model to visually assess performance across different thresholds. Confusion matrices for MLP-TF were also constructed to provide counts of true positives, true negatives, false positives, and false negatives, thereby offering a comprehensive evaluation of classification outcomes.

### Model Explainability

SHAP (*SHapley Additive exPlanations*) value analysis (30) was conducted for the models where applicable, which helped in interpreting the contribution of each feature to the prediction, thus improving the transparency, and understanding of model decisions, which is undoubtedly crucial for clinical applicability and trustworthiness of the models.

## 3.0 Results

### 3.1 Descriptive Statistics

The mean age of patients was 66. Among patients with Merkel cell carcinoma, the majority were male (65.01%), and chemotherapy was given to a small portion (9.33%). At follow-up, most patients (74.64%) were deceased. The majority of tumors were classified as T1 (65.23%), with no lymph node involvement in 74.64% of cases and no distant metastasis in 94.9%. Most patients were married (60.79%) and underwent surgery (93.22%), with 52.7% in AJCC stage 0. Racially, 96.65% were White, and tumors were primarily located on the scalp and neck (23.83%). Tumor laterality was fairly balanced between left and right. Radiation was the most common adjuvant therapy (52.04%), and 46.06% of patients received combined surgery and radiation. Surgical approaches varied, with gross excision (32.87%) being most common (**Table 1**). **Figure 1** shows the overall increasing trend in head and neck MCC diagnoses across the years 2004-2015 in our final sample.

**Figure 1:**
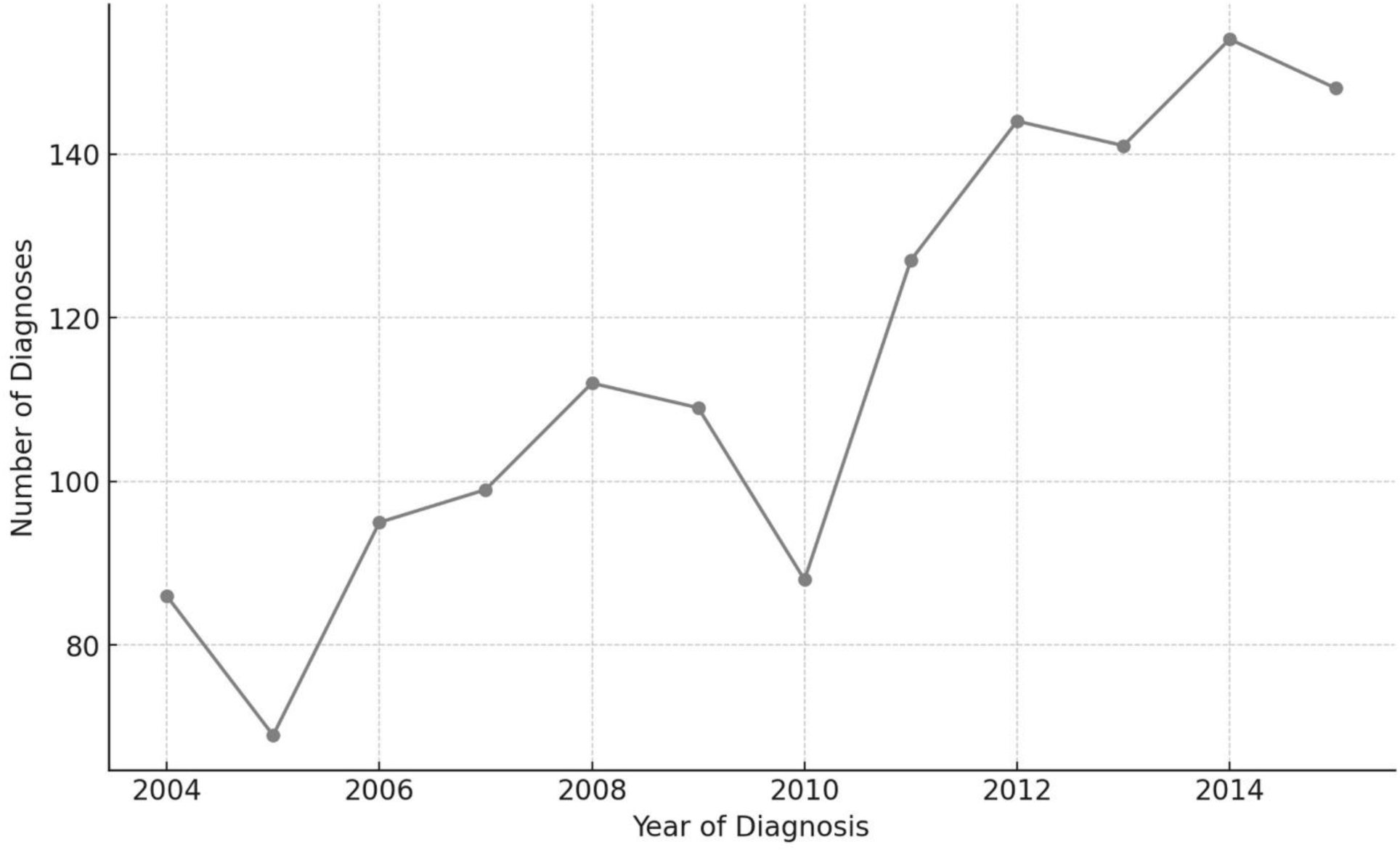
Numbers of head and neck Merkel cell carcinoma diagnoses across the years 2004-2015.

**Table 1:** Descriptive statistics and sample characteristics. Cx: Chemotherapy; Rx: Radiotherapy; NOS: Not Otherwise Specified. **Note:** *Cx and Rx were derived from SEER fields that constituted the categories “Yes” (for Cx) or type of radiation (for Rx) AND “No(ne)/Unknown,” so they may not be 100% indicative of receiving the corresponding therapies since “Unknown” remains a (minor) possibility*.

| **Variable** | **N** | **%** |
| --- | --- | --- |
| Sex |  |  |
| Male | 892 | 65.01 |
| Female | 480 | 34.99 |
| Race |  |  |
| Asian / Pacific Islander | 29 | 2.11 |
| Black | 17 | 1.24 |
| White | 1326 | 96.65 |
| Marital Status |  |  |
| Married | 834 | 60.79 |
| Not Married | 538 | 39.21 |
| T Stage |  |  |
| T0 | 13 | 0.95 |
| T1 | 895 | 65.23 |
| T2 | 290 | 21.14 |
| T3 | 40 | 2.92 |
| T4 | 134 | 9.77 |
| N Status |  |  |
| 0 | 1024 | 74.64 |
| 1 | 348 | 25.36 |
| M Status |  |  |
| 0 | 1302 | 94.9 |
| 1 | 70 | 5.1 |
| AJCC Stage |  |  |
| I | 723 | 52.7 |
| II | 205 | 14.94 |
| III | 374 | 27.26 |
| IV | 70 | 5.1 |
| Primary Site |  |  |
| External ear | 114 | 8.31 |
| Skin lip | 63 | 4.59 |
| Skin of scalp and neck | 327 | 23.83 |
| Other | 868 | 63.27 |
| Tumor Laterality |  |  |
| Left | 550 | 40.09 |
| Not paired site | 295 | 21.5 |
| Paired site midline tumor | 16 | 1.17 |
| Right | 511 | 37.24 |
| Surgery |  |  |
| Performed | 1279 | 93.22 |
| Not Performed | 93 | 6.78 |
| Chemotherapy |  |  |
| No/unknown | 1244 | 90.67 |
| Yes | 128 | 9.33 |
| Adjuvant therapy |  |  |
| None | 638 | 46.5 |
| Chemotherapy | 20 | 1.46 |
| Radiotherapy | 714 | 52.04 |
| Therapies Administered (Overall) |  |  |
| Cx + Rx | 21 | 1.53 |
| Cx only | 8 | 0.58 |
| Rx only | 64 | 4.66 |
| Surgery | 548 | 39.94 |
| Surgery + Cx | 23 | 1.68 |
| Surgery + Rx ± Cx | 708 | 51.6 |
| Surgery Type |  |  |
| No surgery | 93 | 6.78 |
| Local tumor excision, NOS | 286 | 20.85 |
| Gross | 451 | 32.87 |
| Mohs | 139 | 10.13 |
| Wide excision | 403 | 29.37 |

### 3.2 Correlations

Figure 2 shows the correlation matrices for variables of interest in this study. Age, AJCC Stage, and T stage had significant negative Spearman’s and Kendall’s tau-b correlations with overall survival months (p < 0.001) (**Supplementary Table 2**).

**Figure 2:**
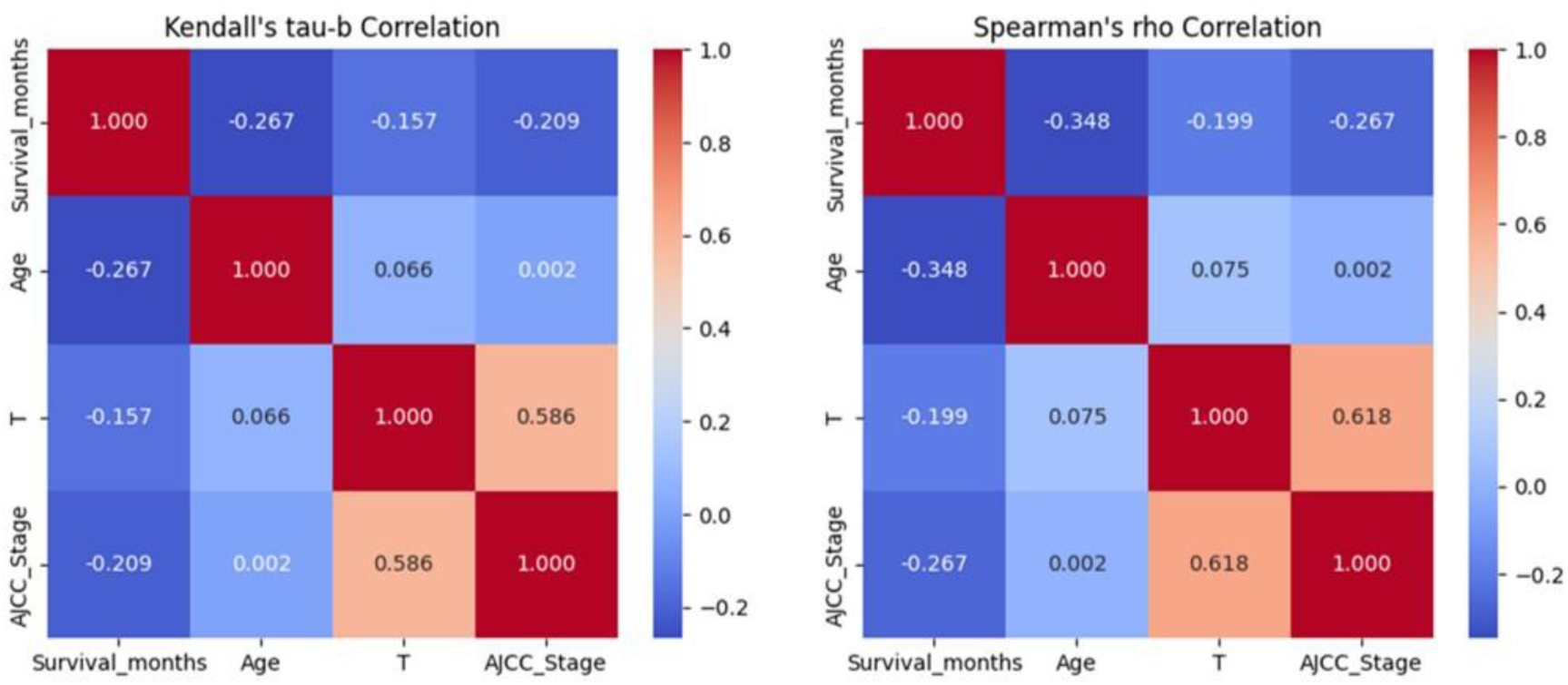
Spearman’s rho and Kendal’s tau-b correlation matrices for ordinal and continuous variables.

### 3.3 Cox Regression and Factors Associated with Overall Survival

The results from the Cox Proportional Hazards model indicated that female patients and those identified Asian/Pacific Islander experienced better survival outcomes in Merkel cell carcinoma. Conversely, older age, higher AJCC stage, the presence of distant metastasis, and having primary tumors located on the scalp and neck were significantly associated with poorer survival rates (Figure 3). Mann-Whitney U tests summarizing difference in overall survival months were consistent (**Supplementary Table 3**).

**Figure 3:**
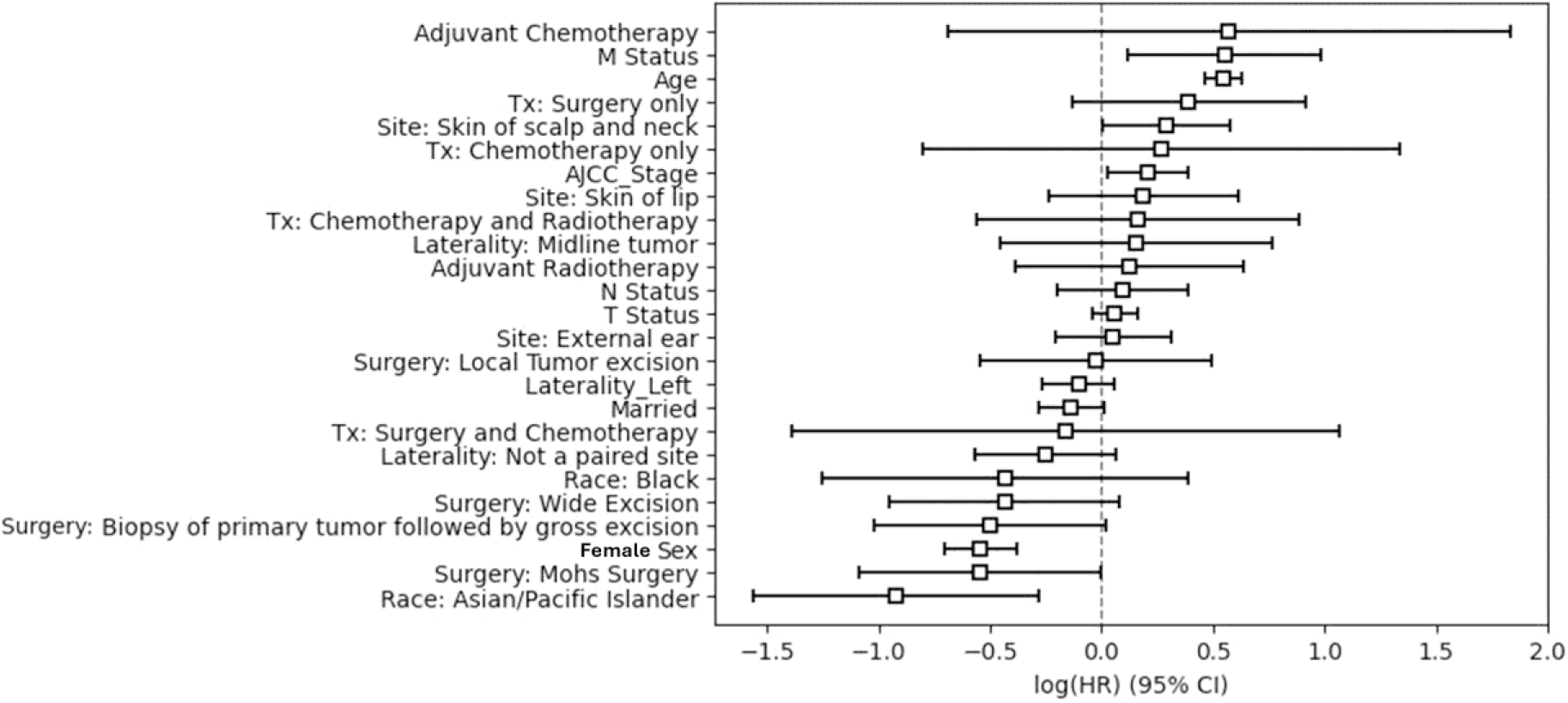
Forest plot summarizing multivariate cox-proportional hazards regression model outcomes for overall survival. Sex: Males (1), Females (0). **Reference categories**: Race (White), Site (Other), Laterality (Right), Adjuvant therapy (absence of such therapy (None)), Therapies (Surgery + Radiotherapy ± Chemotherapy /Radiotherapy only), and Surgery: No Surgery. **Note**: *“Radiotherapy only” was equivalent to not having surgery (0 for all surgery types recoded), “Chemotherapy only” = 0, and “Chemotherapy and Radiotherapy” = 0; “Surgery + Radiotherapy ± Chemotherapy” is equivalent to having surgery (1 for any type of surgery recoded) and “Surgery + Chemotherapy” = 0; ; such variables were excluded to avoid redundancy*.

### 3.4 Model Performance

**Table 2** shows the performance of various models in predicting 5- and 2-year survival; the models were ranked according to their AUROC values. In predicting 5-year survival, the GB model achieved the highest performance with an AUROC of 0.752 (95% CI [0.692, 0.812]), followed by the ADA model with an AUROC of 0.744 (95% CI [0.683, 0.805]), and the LR model ranking third with an AUROC 0.733 (95% CI [0.671, 0.795]). For the 2-year survival prediction, the GB model again outperformed the others with an AUROC of 0.752 (95% CI [0.696, 0.808]). The RF and ADA models ranked second and showed similar AUROCs in the 2-year survival prediction (AUROC = 0.747, 95% CI [0.690, 0.804] and AUROC = 0.747, 95% CI [0.690, 0.804]), while the LR model ranked third (AUROC = 0.740, 95% [0.682, 0.798]). The SVM model had the lowest AUROC for both 5- and 2-year survival predictions (AUROC = 0.624, 95% CI [0.556, 0.692] and AUROC = 0.641, 95% CI [0.576, 0.706], respectively). Models showed higher specificity in 5-year survival predictions compared to 2-year predictions, in general, with the RF model achieving the highest specificity (0.933) for 5-year survival. Recall was generally higher for 2-year predictions, with the RF model attaining the highest recall of 0.946. In terms of accuracy, the GB model led both predictions, with a 2-year accuracy of 0.709 and a 5-year accuracy of 0.680, closely followed by the ADA model. The KNN model had the lowest accuracy for 2-year survival (0.669), while the GNB model had the lowest for 5-year survival (0.596) (**Table 2**).

**Table 2:** Performance of 5-year and 2-year survival classification models.

| **Survival Year** | **Model** | **Accuracy** | **Precision** | **Recall** | **Specificity** | **F1 Score** | **AUROC [95% CI]** |
| --- | --- | --- | --- | --- | --- | --- | --- |
| 5 Year | GB | 0.680 | 0.671 | 0.420 | 0.859 | 0.516 | 0.752 [0.692, 0.812] |
|  | ADA | 0.680 | 0.667 | 0.429 | 0.853 | 0.522 | 0.744 [0.683, 0.805] |
|  | LR | 0.673 | 0.667 | 0.393 | 0.865 | 0.494 | 0.733 [0.671, 0.795] |
|  | MLP-S5 | 0.658 | 0.636 | 0.375 | 0.853 | 0.472 | 0.729 [0.667, 0.791] |
|  | TF-MLP | 0.662 | 0.564 | 0.750 | 0.601 | 0.644 | 0.729 [0.667, 0.791] |
|  | RF | 0.647 | 0.703 | 0.232 | 0.933 | 0.349 | 0.726 [0.664, 0.788] |
|  | KNN | 0.644 | 0.621 | 0.321 | 0.865 | 0.424 | 0.713 [0.650, 0.776] |
|  | DT | 0.636 | 0.586 | 0.366 | 0.822 | 0.451 | 0.670 [0.604, 0.736] |
|  | GNB | 0.596 | 0.503 | 0.688 | 0.534 | 0.581 | 0.667 [0.601, 0.733] |
|  | SVM | 0.669 | 0.662 | 0.384 | 0.865 | 0.486 | 0.624 [0.556, 0.692] |
| 2 Year | GB | 0.709 | 0.744 | 0.798 | 0.570 | 0.770 | 0.752 [0.696, 0.808] |
|  | RF | 0.716 | 0.697 | 0.946 | 0.355 | 0.803 | 0.747 [0.690, 0.804] |
|  | ADA | 0.709 | 0.744 | 0.798 | 0.570 | 0.770 | 0.747 [0.690, 0.804] |
|  | LR | 0.702 | 0.729 | 0.815 | 0.523 | 0.770 | 0.740 [0.682, 0.798] |
|  | MLP-S2 | 0.691 | 0.734 | 0.774 | 0.561 | 0.754 | 0.736 [0.678, 0.794] |
|  | TF-MLP | 0.709 | 0.722 | 0.851 | 0.486 | 0.781 | 0.729 [0.670, 0.788] |
|  | DT | 0.705 | 0.706 | 0.887 | 0.421 | 0.786 | 0.727 [0.668, 0.786] |
|  | KNN | 0.669 | 0.693 | 0.821 | 0.430 | 0.752 | 0.716 [0.656, 0.776] |
|  | GNB | 0.673 | 0.689 | 0.845 | 0.402 | 0.759 | 0.693 [0.631, 0.755] |
|  | SVM | 0.698 | 0.696 | 0.899 | 0.383 | 0.784 | 0.641 [0.576, 0.706] |

MLP-S and TF-MLP showed similar recall scores in predicting the 2-year survival (0.774 and 0.851, respectively), however, marked differences in the 5-year recall scores were shown between the two models (0.375 and 0.750, respectively). The accuracy of the models was comparable across both the 2-year and 5-year survival prediction groups (2-year survival accuracy: MLP-S = 0.691, TF-MLP = 0.709; 5-year survival accuracy: MLP-S = 0.658, TF-MLP = 0.662). Moreover, the specificity for 2-year survival predictions was lower in both models (MLP-S = 0.561, TF-MLP = 0.486) compared to the specificity observed in the 5-year survival predictions (MLP-S = 0.853, TF-MLP = 0.601) (**Table 2**).

**Table 3** summarizes the performance of regression models in predicting overall survival months. Ridge regression performed best, with an MAE of 939.978, an MSE of 1,399,111.691, an RMSE of 1,182.841, and an R² of 0.226, while Lasso regression followed closely with an R² of 0.223. Ensemble models like Gradient Boosting (GB) and Random Forest (RF) had R² values of 0.217 and 0.184, respectively, but did not surpass simpler linear models. Meanwhile, the Multi-Layer Perceptron (MLP) Regressor and Support Vector Regressor (SVR) had lower predictive power, reflected in higher errors and lower R² values (0.167 and 0.166, respectively).

**Table 3:** Performance of overall survival months regression models. MAE: Mean Absolute Error; MSE: Mean Squared Error; RMSE: Root Mean Squared Error; R^2^: Coefficient of Determination.

| **Model** | **MAE** | **MSE** | **RMSE** | **R<sup>2</sup>** |
| --- | --- | --- | --- | --- |
| Linear Regression | 942.357 | 1420727.551 | 1191.943 | 0.214 |
| MLP Regressor | 963.364 | 1506051.306 | 1227.213 | 0.167 |
| Gradient Boosting Regressor | 951.055 | 1415232.796 | 1189.636 | 0.217 |
| Random Forest Regressor | 985.815 | 1474160.170 | 1214.150 | 0.184 |
| Ridge | 939.978 | 1399111.691 | 1182.841 | 0.226 |
| Lasso | 945.362 | 1403529.308 | 1184.706 | 0.223 |
| SVR | 943.494 | 1508186.815 | 1228.083 | 0.166 |

For F1 scores, they were generally higher in 2-year survival prediction compared with 5-year (**Table 2**). The TF-MLP model excelled in 5-year survival (0.644), ranking fourth for 2-year predictions (0.781). The RF model achieved the highest F1 score (0.803) for 2-year predictions, but it ranked lowest in 5-year predictions (0.349) (**Table 2**). The TF-MLP model reached an AUROC of 0.729 for both survival predictions (Figure 4a), with confusion matrices showing 52 true negatives, 55 false positives, 25 false negatives, and 143 true positives for the 2-year model, and 98 true negatives, 65 false positives, 28 false negatives, and 84 true positives for the 5-year model (Figure 4b). ROC curves for top three performing models are shown in Figure 4c.

**Figure 4:**
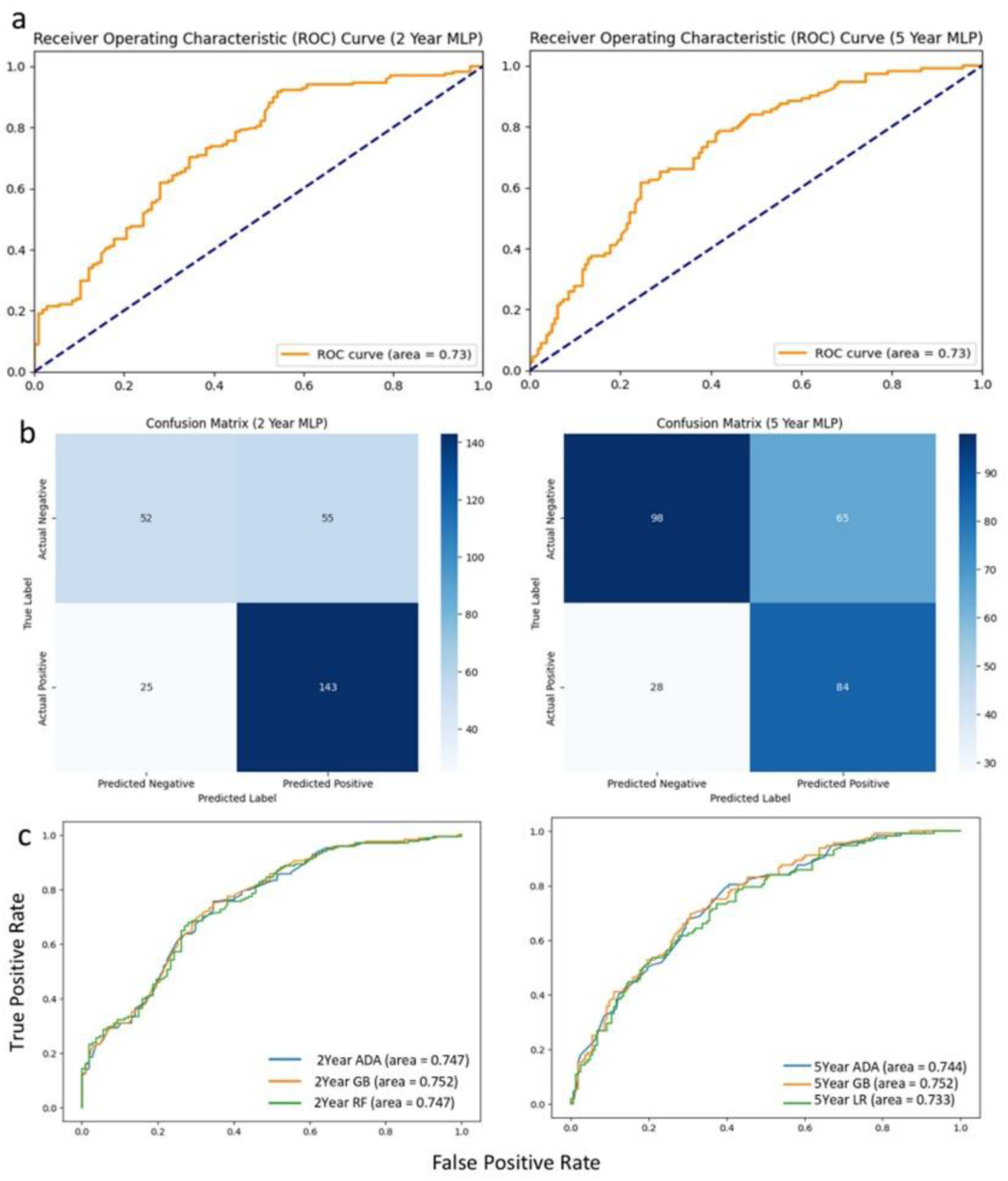
(a) ROC curves, and (b) confusion matrices for TF-MLP. (c) ROC curves for top three models, 2-year survival (left) and 5-year survival (right).

Survival model performance was also evaluated using the C-index, with the Cox Proportional Hazards (CPH) model achieving the highest score (0.7), followed by the Random Forest Survival (RFS) model (0.591) and the Survival Tree model (0.582) (**Table 4**).

**Table 4:** C-Indices of survival models. RFS: Random Forest Survival; CPH: Cox-Proportional Hazards Regression.

| **Model** | **C-Index** |
| --- | --- |
| RFS | 0.591 |
| Survival<br>Tree | 0.582 |
| CPH | 0.700 |

### 3.5 SHAP Analysis

Figure 5 shows the SHAP plots depicting the contribution of predictors in top-performing classification (GB) and regression (Ridge Regression) models. Age, sex, and AJCC stage were the top three influential features in both models.

**Figure 5:**
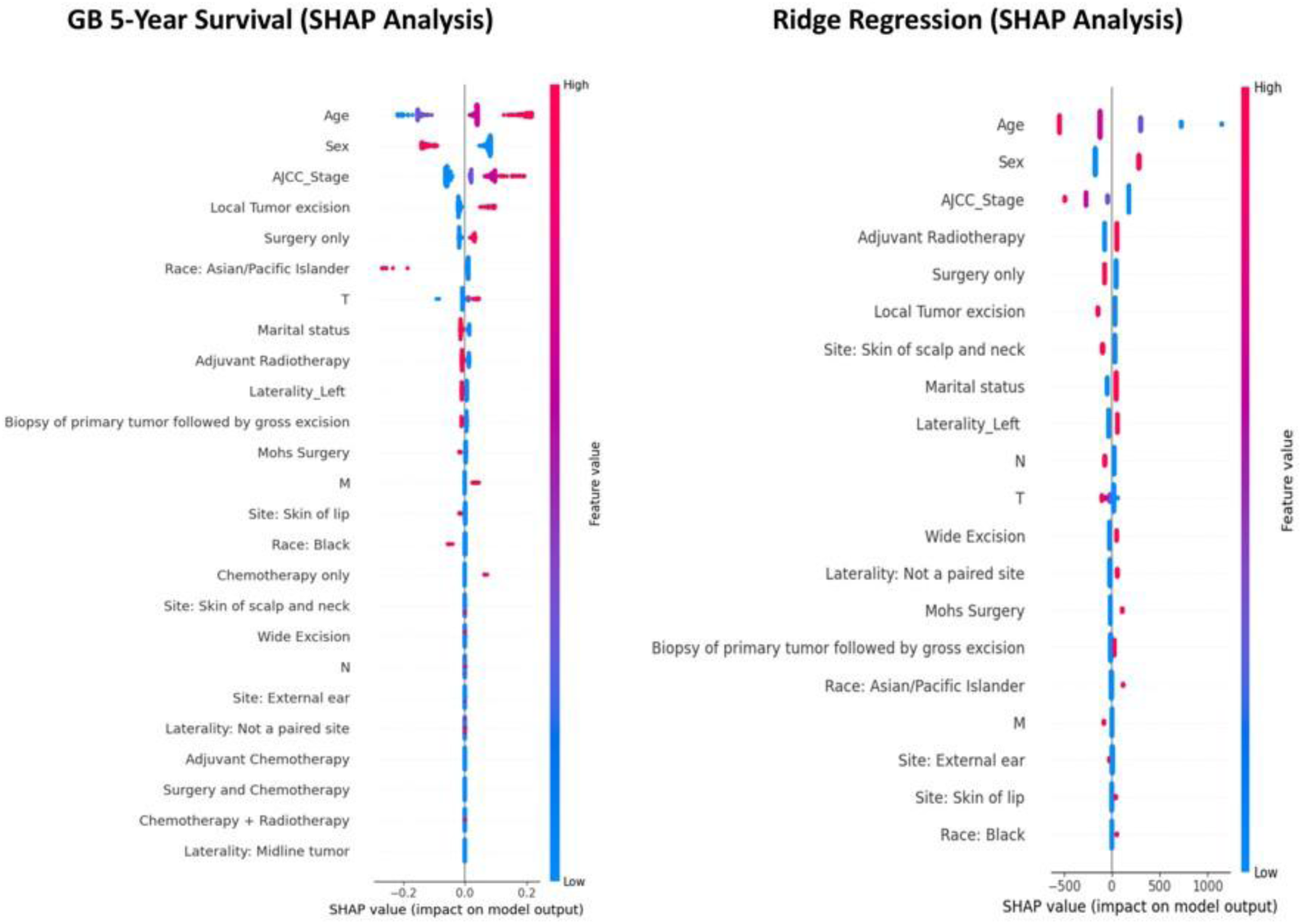
SHAP plot for top performing classification (left) and regression (right) models.

## 4.0 Discussion

Predicting the prognosis of Merkel cell carcinoma (MCC) remains a challenge due to its aggressive nature and complex biological behavior (31).This study aimed to evaluate the predictive performance of various models in forecasting survival outcomes for patients with MCC. Across the models, the 2-year survival prediction was more accurate than the 5-year survival group, overall. Some machine learning models, such as best-performing model in thus study, GB, had acceptable predictions in both durations, while SVM algorithm performed the worst.

According to our results, older patients, males, and those with advanced AJCC stages demonstrated significantly worse survival outcomes, consistent with findings from previous research. Age has been shown to be a major factor affecting MCC survival, with older patients experiencing poorer outcomes (32, 33). Additionally, our results reveal that male patients with MCC experience worse survival outcomes than female patients, a trend observed in several cancers (33, 34). AJCC staging remains a crucial predictor of survival in MCC, as numerous studies have demonstrated that patients with advanced-stage tumors, particularly those with nodal involvement or distant metastases, face significantly lower survival rates (35).

Lasso and Ridge are regularization techniques applied to linear regression models, widely used in cancer prognosis for their ability to handle high-dimensional data and effectively select variables for model construction (36, 37). They have shown strong predictive performance in cancers such as head and neck squamous cell carcinoma when combined with clinical characteristics (38). In our study, Ridge and Lasso outperformed RF in predicting survival months, consistent with findings that Lasso-type models can surpass more complex approaches like GB, which may sometimes have higher false-positive rates (39).

The Cox Proportional Hazards (CPH) model is a multivariate statistical tool for survival analysis. It provides hazard ratios for variables, aiding clinicians in interpreting results and making decisions. Understanding its rationale and assumptions is essential for its effective application in time-to-event studies (40, 41, 42). In contrast, RFS models extends random forests to handle right-censored survival data, effectively managing multiple covariates, noise, and complex nonlinear relationships without prior specifications. (43, 44). While survival trees serve as a flexible, nonparametric alternative to parametric models, automatically detecting interactions without prearrangement (43). Through an analysis of the performance of survival models, CPH, RFS, and Survival Trees, the CPH model was identified as having the best discrimination, with a C-index of 0.7, making it valuable in clinical prognostic settings. The RFS and Survival Tree models for MCC yielded lower C-index values of 0.591 and 0.582, respectively. However, this was in contrast to a study on oral squamous cell carcinoma, where RFS outperformed CPH in survival predictions (45). On the other hand, a study on laryngeal squamous cell carcinoma found that the CPH model achieved a moderate C-index of 0.747, while RFS showed a lower C-index of 0.596 (46). These findings further support the need for incorporating additional clinical factors, as AI models like Deep Neural Networks have shown significantly higher prediction accuracy in such contexts.

In our survival prediction analysis, the GB classification model demonstrated strongest performance with an AUC value of 0.752 (95% CI: 0.696-0.808) and 0.752 (95% CI: 0.692-0.8120 for both 2-year and 5-year survival predictions, respectively, outperforming all other models. This aligns with findings in a colorectal cancer survival study, where the GB model also performed well but with a higher AUC of 0.867 for 5-year predictions (47). On the other hand, the LR model ranked higher than all other models in the colorectal cancer study with an AUC of 0.872, while in our analysis it came third after the ADA and GB models with an AUC of 0.733. GB models have been successfully used for their ability to handle nonlinear interactions between variables (48). It was also found that Gradient boosting machine outperforms traditional methods like AJCC staging in predicting cancer-specific survival for resected intrahepatic cholangiocarcinoma (49).

According to our results, RF also showcased acceptable predictions in the 2-year survival of patients (AUROC = 0.747, 95% CI [0.690, 0.804]), although it ranked sixth among other models in the 5-year survival group (AUROC = 0.726 [0.664, 0.788]). However, the RF model has shown to be effective in selecting genomic biomarkers, especially for multi-class tasks (50, 51). In a study using data from head and neck non-squamous cell carcinoma patients to predict post-treatment survival (52), RF had higher AUC values than regression prediction with TNM stages, in which RF predicted 5-year overall survival with an AUROC of 0.862 (95% CI [0.842, 0.882]).

A previous study about breast cancer identified ADA as one of the most effective predictors for survival, where in the 5-year survival analysis it showcased an AUC value of 0.779, supporting the strength of our results, which yielded an AUROC of 0.744 (95% CI [0.683, 0.805]). Furthermore, LR model directly ranked behind ADA across both Breast cancer and MCC with an AUC of 0.768 and 0.733 respectively. Together, these results demonstrate ADA’s leading role in survival prediction, with LR coming out as a reliable and potential competitor (53).

SVM shows strong results in early cancer detection such as pancreatic cancer (54), but it underperformed in predicting MCC survival, where it ranked worst in survival predictability among both the 5 and 2-year groups (AUROC = 0.624; 95% CI [0.556, 0.692] and AUROC = 0.641; 95% CI [0.576, 0.706], respectively). Although, SVM matched RF in breast cancer survival predictions (55).

To ensure explainability of our AI model prediction, our SHAP analysis, powered by a game-theoretic interpretability approach, was done for both the GB and Ridge regression models. Influential predictors aligned closely with our findings on CPH-derived survival predictors in MCC. SHAP highlights age, sex, and AJCC stage as the most significant variables influencing model survival prediction, further confirming that older patients, males, and those with advanced AJCC stages face significantly worse survival rates (56).

### Limitations

Despite the promising results, our study has several limitations. First, the dataset, while comprehensive, primarily included White patients, limiting the generalizability of our findings to other racial groups. Future studies should include larger and more diverse populations to validate these findings and examine the effects of various treatment modalities on survival outcomes. Additionally, our analysis was restricted to clinicodemographic predictors, which may have omitted important radiomic and molecular features that could enhance prediction accuracy. Some variables like receiving chemotherapy or radiotherapy were not definitive and had unknowns, raising a limitation. Further studies should explore the inclusion of radio-genomic data and evaluate progression-free survival (PFS) alongside overall survival to provide a more comprehensive assessment. Lastly, while we tested a broad range of machine learning models, including neural networks, the relatively small sample size may have constrained their performance.

## Conclusion

This study assessed various machine learning models to predict overall survival outcomes for patients with Merkel Cell Carcinoma. Significant predictors identified were age, sex, and AJCC stage. Older age, male sex, and advanced stages were associated with poorer survival, aligning with existing MCC research. Across different tested models, the Gradient Boosting model emerged as the top-performing for both 2-year and 5-year overall survival predictions. Ridge-regularized regression performed well in predicting overall survival months. Additionally, SHAP analysis enhanced model interpretability by confirming that key clinical variables significantly influenced survival predictions. Future studies should focus on validating these models in different populations and incorporate additional clinical and molecular data to further enhance predictive performance and support personalized patient care.

## Author Contributions

Jehad A. Yasin and Abd-alrahman Obeid: Conceptualization, Methodology, Formal Analysis, Data Curation, and Writing – Original Draft preparation; Fares A. Qtaishat: Methodology and Writing – Original Draft preparation; Hanan M. Qasem: Writing – Original Draft; Mohammad-Amer A. Tamimi and Sakher Alshwayyat: Investigation and Resources, Writing – Original Draft preparation; Osama M. Younis: Data Curation, Formal Analysis, Writing – Review & Editing; Yousef A. Ateiwi: Writing – Review & Editing;, and contributed to Writing – Review & Editing. Nicole Issi: Investigation, Visualization; Ramez M. Odat: Conceptualization, Supervision, Project Administration, and Writing – Review & Editing.

## Data Availability Statement

The data generated in this study are available upon request from the corresponding author.

## Acknowledgements

None.

## Ethical Considerations

No ethical approval was required; research was done on open-access data.

## Supplementary Material

**Supplementary Table 1:**
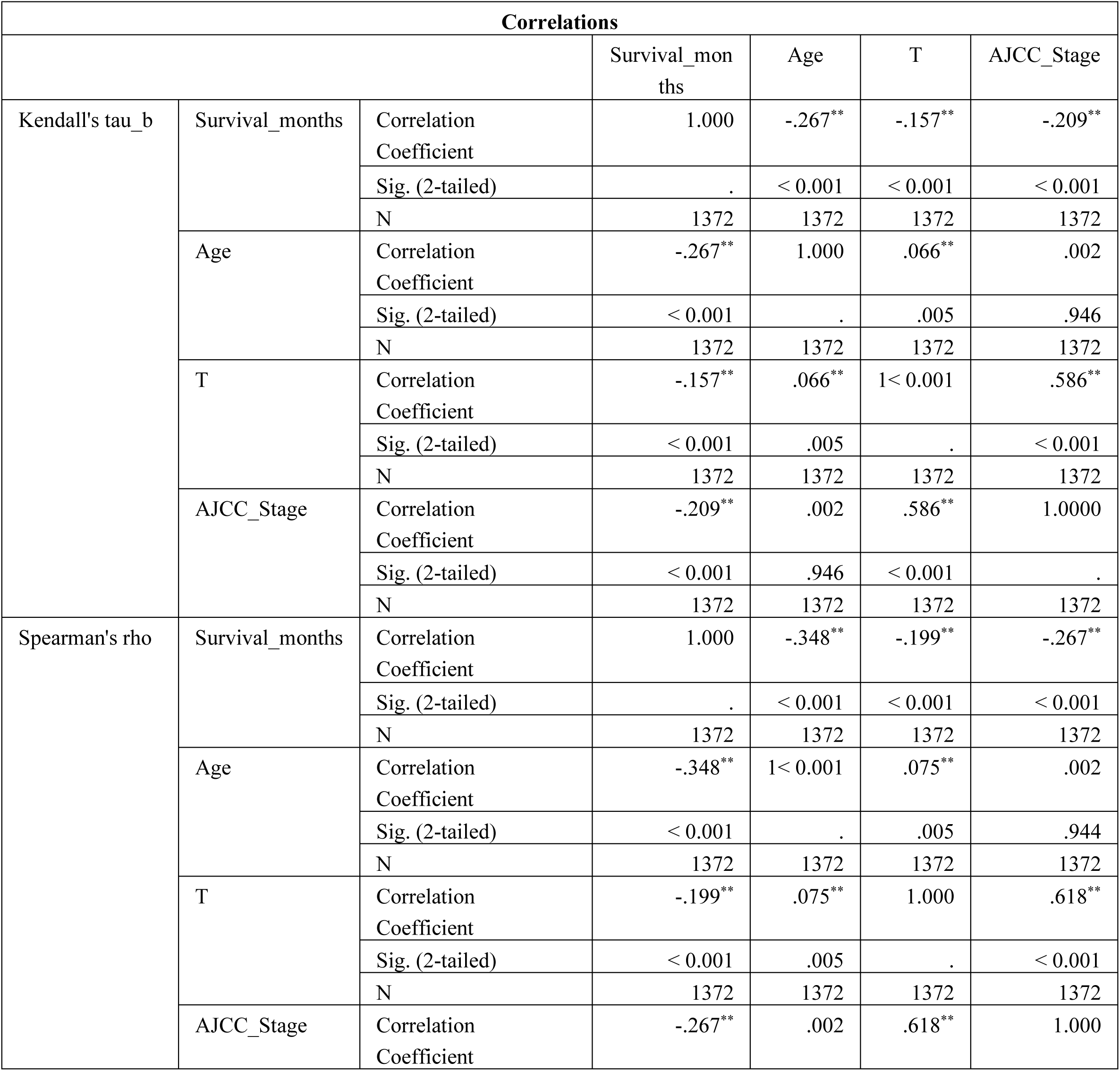
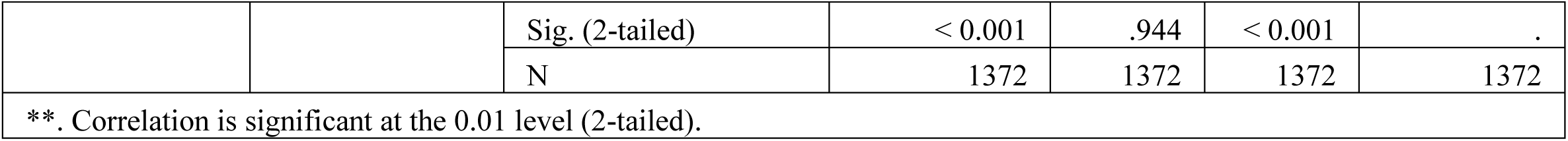
Correlations between variables of interest.

**Supplementary Table 2:**
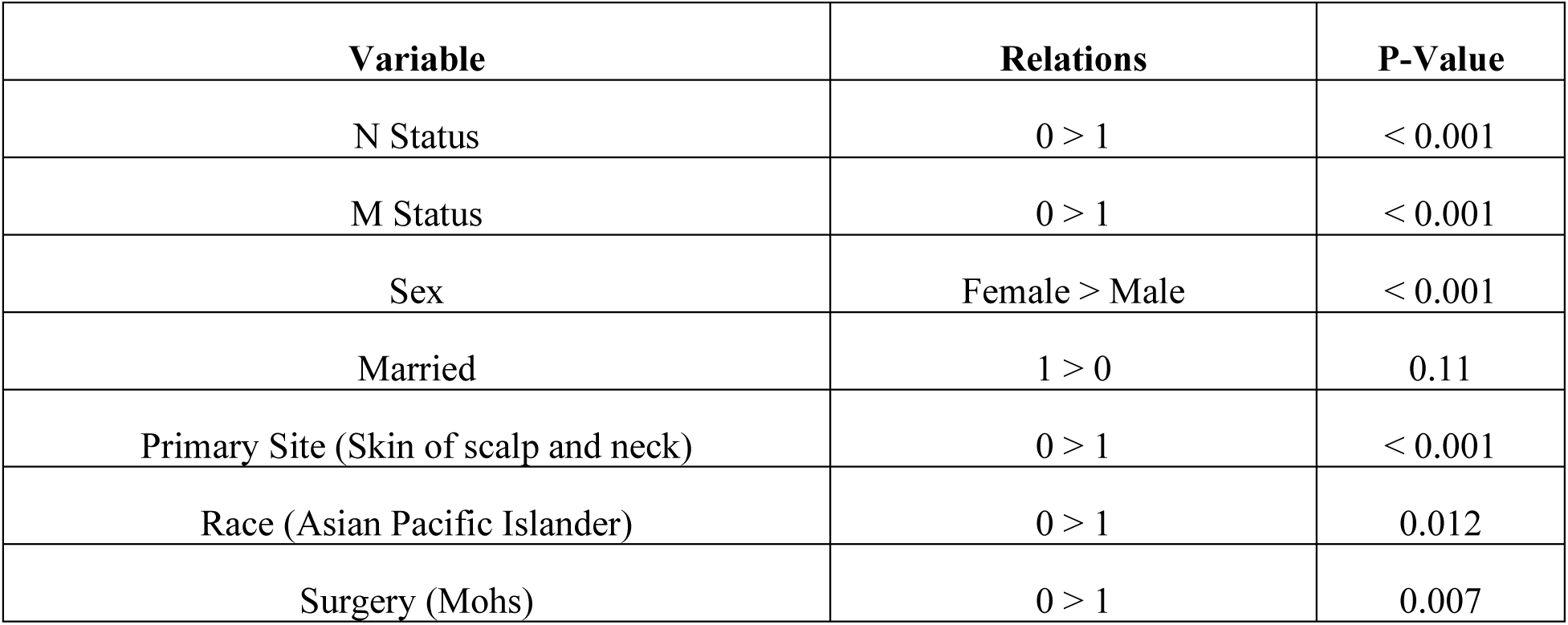
Overall survival months across different binary variables. Test of significance: Mann-Whitney U Test; 0: No, 1: Yes.

| Variable | Relations | P-Value |
| --- | --- | --- |
| N Status | 0 > 1 | < 0.001 |
| M Status | 0 > 1 | < 0.001 |
| Sex | Female > Male | < 0.001 |
| Married | 1 > 0 | 0.11 |
| Primary Site (Skin of scalp and neck) | 0 > 1 | < 0.001 |
| Race (Asian Pacific Islander) | 0 > 1 | 0.012 |
| Surgery (Mohs) | 0 > 1 | 0.007 |

**Supplementary Table 3:** Best parameters for classification models constructed using scikit-learn. MLPS2 has two hidden layers, while MLPS5 has one hidden layer.

| Model | Hyperparameters |
| --- | --- |
| **2yearKNN** | leaf_size=20, metric='euclidean', n_neighbors=101, p=1, weights='uniform' |
| **5yearKNN** | leaf_size=20, metric='euclidean', n_neighbors=95, p=1, weights='uniform' |
| **2yearSVM** | C=0.1, gamma='scale', kernel='poly' |
| **5yearSVM** | C=0.05, gamma='scale', kernel='linear', probability=True |
| **2yearLR** | C=0.1, penalty='l1', solver='saga' |
| **5yearLR** | C=0.1, penalty='l2', solver='saga' |
| **2yearDT** | max_depth=3, min_samples_leaf=10, min_samples_split=2 |
| **5yearDT** | max_depth=10, min_samples_leaf=10, min_samples_split=2 |
| **2yearRF** | max_depth=3, min_samples_split=2, n_estimators=500 |
| **5yearRF** | max_depth=3, min_samples_split=40, n_estimators=250 |
| **2yearGB** | learning_rate=0.2, max_depth=1, n_estimators=50 |
| **5yearGB** | learning_rate=0.1, max_depth=1, n_estimators=200 |
| **2yearADA** | learning_rate=0.1, n_estimators=100 |
| **5yearADA** | learning_rate=1, n_estimators=10 |
| **2yearGNB** | var_smoothing=1e-120 |
| **5yearGNB** | var_smoothing=1e-30 |
| **2yearMLP (MLPS2)** | activation='relu', alpha=0.05, hidden_layer_sizes=(50, 50), learning_rate='constant', learning_rate_init=0.001, solver='sgd' |
| **5yearMLP (MLPS5)** | activation='relu', alpha=0.05, hidden_layer_sizes=(100,), learning_rate='adaptive', learning_rate_init=0.001, solver='sgd' |

